# Generative Methods for Pediatric Genetics Education

**DOI:** 10.1101/2023.08.01.23293506

**Authors:** Rebekah L. Waikel, Amna A. Othman, Tanviben Patel, Suzanna Ledgister Hanchard, Ping Hu, Cedrik Tekendo-Ngongang, Dat Duong, Benjamin D. Solomon

## Abstract

Artificial intelligence (AI) is used in an increasing number of areas, with recent interest in generative AI, such as using ChatGPT to generate programming code or DALL-E to make illustrations. We describe the use of generative AI in medical education. Specifically, we sought to determine whether generative AI could help train pediatric residents to better recognize genetic conditions. From publicly available images of individuals with genetic conditions, we used generative AI methods to create new images, which were checked for accuracy with an external classifier. We selected two conditions for study, Kabuki (KS) and Noonan (NS) syndromes, which are clinically important conditions that pediatricians may encounter. In this study, pediatric residents completed 208 surveys, where they each classified 20 images following exposure to one of 4 possible educational interventions, including with and without generative AI methods. Overall, we find that generative images perform similarly but appear to be slightly less helpful than real images. Most participants reported that images were useful, although real images were felt to be more helpful. We conclude that generative AI images may serve as an adjunctive educational tool, particularly for less familiar conditions, such as KS.

## Introduction

Deep learning (DL) is a sub-field of artificial intelligence (AI) that uses multiple neural network layers to analyze data.^1^ DL has become a powerful tool adopted across a wide range of biomedical research disciplines and has strong clinical potential. In medical settings, DL has been used to perform tasks such as identifying which individuals might be at risk of poor outcomes, generating a differential diagnosis based on images such as radiologic or pathologic studies, and recommending treatment options.^1–3^ In genomics, DL and related methods can be used to help analyze DNA sequences or other laboratory-based information as well as phenotypic data.^2–4^ Deep learning (DL) is a sub-field of artificial intelligence (AI) that uses multiple neural network layers to analyze data.^1^ DL has become a powerful tool adopted across a wide range of biomedical research disciplines and has strong clinical potential. In medical settings, DL has been used to perform tasks such as identifying which individuals might be at risk of poor outcomes, generating a differential diagnosis based on images such as radiologic or pathologic studies, and recommending treatment options.^1–3^ In genomics, DL and related methods can be used to help analyze DNA sequences or other laboratory-based information as well as phenotypic data.^2–5^

Generative AI is a relatively new branch of AI in which new data can be created through training on existing datasets. One type of generative AI that can be used to create images uses “generative adversarial networks” (GANs).^6^ These generated images can be used for different purposes, such as to enhance small datasets and increase the diversity of available images.^7, 8^ Another possible use of generative AI is for medical education.^9, 10^ For example, many fake images can be quickly created and customized to help expose radiologist or pathologist trainees to a large number of X-ray, MRI, or hematologic images.^11^

These types of AI technologies can be especially advantageous in the field of rare genetic diseases, as they can enable increased exposure to images of individuals with very rare conditions and greater recognition of dysmorphology in various populations.^12^ This is particularly salient because, despite the high proportion of pediatric individuals with genetic conditions,^13, 14^ pediatric residents may have sparse exposure to medical genetics training and insufficient educational resources, including the lack of standardized training requirements in medical genetics by Accreditation Council for Graduate Medical Education (ACGME). (https://www.acgme.org/globalassets/pfassets/programrequirements/320_pediatrics_2022_tcc.pdf). This issue, compounded by the shortage of medical geneticists, necessitates creative solutions to help optimize early diagnosis and management of individuals affected by genetic conditions.^15^

Despite the lack of standardized genetics training in pediatric residencies, some programs have implemented innovative strategies to support genetic competencies. These include immersive learning methods such as personal genome sequencing, interdisciplinary and interprofessional education through cross-specialty training, didactic courses, workshops, or alternative electronic-learning methods. Such approaches may be particularly beneficial for programs that have limited contact with medical geneticists, or where a genetics elective may not be feasible.^16, 17^ In one survey, 87% of pediatric residents agreed that an online-based module could effectively deliver genetics education,^18^ and an email-based approach was helpful and cost-effective in disseminating novel genomic information to physicians.^19^ Additionally, a recent survey found medical students have a high interest and desire to learn about AI in medicine.^19, 20^

In the spirit of these innovations, we conducted a study to investigate the potential use of generative AI in training pediatric residents to identify specific genetic conditions.

## Materials and Methods

### Data collection and Image selection

Similar to our previous work,^21, 22^ we identified and used publicly available images depicting individuals with Kabuki syndrome (KS), (OMIM #147920 and #300867)^23^ or Noonan syndrome (NS), (OMIM #163950, 605275, 609942, 610733, 611553, 613706, 615355, 616559, 616564, 619087).^24^ We chose these conditions because they are relatively common genetic disorders with recognizable facial features that can be important to diagnose early due to their clinically relevant but often occult impact on organ systems, such as cardiac anomalies, bleeding diatheses, or oncologic risks, depending on the specific condition.^23, 24^ We also felt that NS may be better known and more recognizable to pediatric residents than KS, and wanted to contrast results.

From the available source information for each image, we documented age, gender, and ancestry, intentionally aiming for diverse representation in these three areas (See File 1). In total, we collected 278 NS images and 239 KS images. Image sets included both color and black-and-white images, with varying image resolution. As we focused on pediatric residents, only pediatric (newborn to ∼18 years old) facial images were used for survey images. To ensure the accuracy of the images used, we checked that the facial images (both real and GAN-based, as described below) used in the survey and in the educational interventions were correctly classified as the syndrome of interest using Face2Gene (https://www.face2gene.com/).^5^

### Generative adversarial network (GAN) generation

Our image generator is based on Nvidia StyleGAN2-ADA. StyleGAN2-ADA creates a fake random image by converting a random vector into this image,^6^ and a label vector (default size 512×1), if available, can also be used with this random vector, thus providing the user more control for generating images with certain criteria (e.g., face images of only young individuals). The default Nvidia StyleGAN2-ADA trained on FFHQ dataset did not use labels. We followed the same approach as in our previous work,^22^ and finetuned the default StyleGAN2-ADA on our own labeled datasets. Our label vector of size 512 consists of three subparts: a vector of size 256 indicating the disease label, a vector of size 128 indicating the age group, and a vector of size 128 indicating the gender. Hence, the additional parameters to be trained are the disease embedding 256 x 11 (for 10 genetic conditions and one unaffected group used for training (see: https://github.com/datduong/stylegan3-syndromic-faces)), the age embedding 128 x 5 for five age groups (under 2 years old, 2-9 years old, 10-19 years old, 20-34 years old, and >35 years old), and gender embedding 128 x 2 for female and male. Due to low sample sizes, we could only train a small number of embeddings. Hence, the age and gender embedding are shared across all images; that is, these embeddings are not specific to a particular genetic condition.

Although we evaluated the GAN application on just KS and NS, we used images of the other conditions during fine-tuning (File 1). Since people with some of these conditions can have similar facial features, using images of people with additional conditions allows us to generate more specific images of people with the genetic conditions of interest.

Because our dataset is small, we fine-tuned Nvidia StyleGAN2-ADA of images size 256 x 256 instead of the higher resolution 1024 x 1024. Following Nvidia image pre-processing, we aligned all the faces so that all images have roughly similar head sizes/orientations and eye positions. We further removed the background by setting these pixel values to zero; this approach helps remove some potential GAN artifacts such as artifactual strands of hair or backgrounds. Since we have different numbers of images of each condition, during fine-tuning we applied sample weighting so that images from each disease have roughly uniform weights (see: https://github.com/datduong/stylegan3-syndromic-faces).

We fine-tuned StyleGAN2-ADA on our images together with their corresponding labels (File 2*). These labels provide more control to generate fake images of a person with a particular genetic condition, age, and gender, and, for the “transformation strips” (see below), allow us to alter these fake images to resemble unaffected individuals by manipulating the disease embeddings. We generated single images as well as what we called “transformation strips”, which depicted an individual changing from unaffected to affected. This was inspired by previous work we had done using GANs to depict disease progression, such as cutaneous findings in neurofibromatosis type 1.^21^ In preliminary testing, medical genetics residents, reported transformation strips to be helpful to identify key features in genetic conditions.

After fine-tuning, we picked a random vector and the label embeddings to generate a random image of a person representing a selected genetic condition, age group, and gender. To be more specific, such as to generate an image of young female individual with KS (and likewise NS), we would provide StyleGAN2-ADA with a random vector and the label embeddings of KS, young child, and female. To make this generated individual with KS look like a person without this condition (for the transformation strips described above), we interpolated the embeddings of KS and unaffected, while keeping the corresponding random vector, the age, and gender embedding unchanged.

### Comparison of Educational Interventions

We compared educational interventions via surveys sent using Qualtrics (Provo, Utah, United States). Surveys were specific to either KS or NS. We compared four different interventions to assess the efficacy of various educational approaches. Survey arms included: 1) text description of facial features (control), 2) text description plus 5 images of real individuals with the genetic condition, 3) text description plus 5 GAN images of the genetic condition, 4) text description plus 5 GAN transformation strips (File 3*). In addition to checking images through an external classifier as described above, clinicians in our research group manually reviewed the images to ensure that they were representative of the condition and did not represent extremes of difficulty.

Following the educational intervention, participants were asked to classify 20 images, as well as rate their confidence level for each classification. Additionally, participants were asked demographic questions and pre– and post-intervention opinion questions about diagnostic facial features and the impact of age, gender, and ancestry on facial diagnostics. Example surveys can be found at https://github.com/datduong/stylegan3-syndromic-faces.

Participants were recruited via email. To identify survey respondents, we obtained email addresses through professional networks, departmental websites, journal publications, and other publicly available lists. A survey was considered complete if all classification questions were answered.

The study was formally approved as IRB exempt by the NIH IRB (IRB# 001115).

In this paper, we include mentions of the figures we prepared; as preprint servers may understandably not allow such images to be shown, even when previously published, we include an asterisk (*) by each figure number to explain why it is not available in this version, or why a portion of the figure is not shown.

## Results

### Comparison of education interventions

Of the 2515 pediatric residents contacted from 40 programs across the country, 208 surveys were completed (see details in Supplemental Files 4 and 5). In the survey, pediatric resident participants were provided either a text description of the genetic condition of interest or a text description along with real or GAN-based images. Following the intervention, the participants classified 20 images as either showing a person with the syndrome of interest (KS or NS) or other syndrome. See File 6 for a comparison of the efficacies of the different interventions including images of other conditions. To determine whether the image interventions improved accuracy classifying the syndrome of interest, we calculated only accuracies of KS or NS images. Baseline accuracies for NS (65.3%) were higher than KS (48.2%) for text description only. All image interventions provided some improvement in accuracy, particularly for KS images (Figure 1), with a 12.1% improvement between text only and text plus real images (p=0.0325). We observed a similar improvement with transformation strips (11.4% improvement, p=0.0315) and a single set of fake images (8.8%, p=0.051). Accuracies for NS images with the addition of real images, GAN images, and transformation strips were 74.3% (p=0.025), 68.0% (p=0.276), and 71.0% (p=0.152), respectively. The p-values allow us to compare the relative effectiveness of these experiments; for example, real, GAN, and transformation images have much lower p-values in KS than NS, thus real images and GAN-based images do provide improvement over text-only in KS, but not in NS.

**Figure 1.**
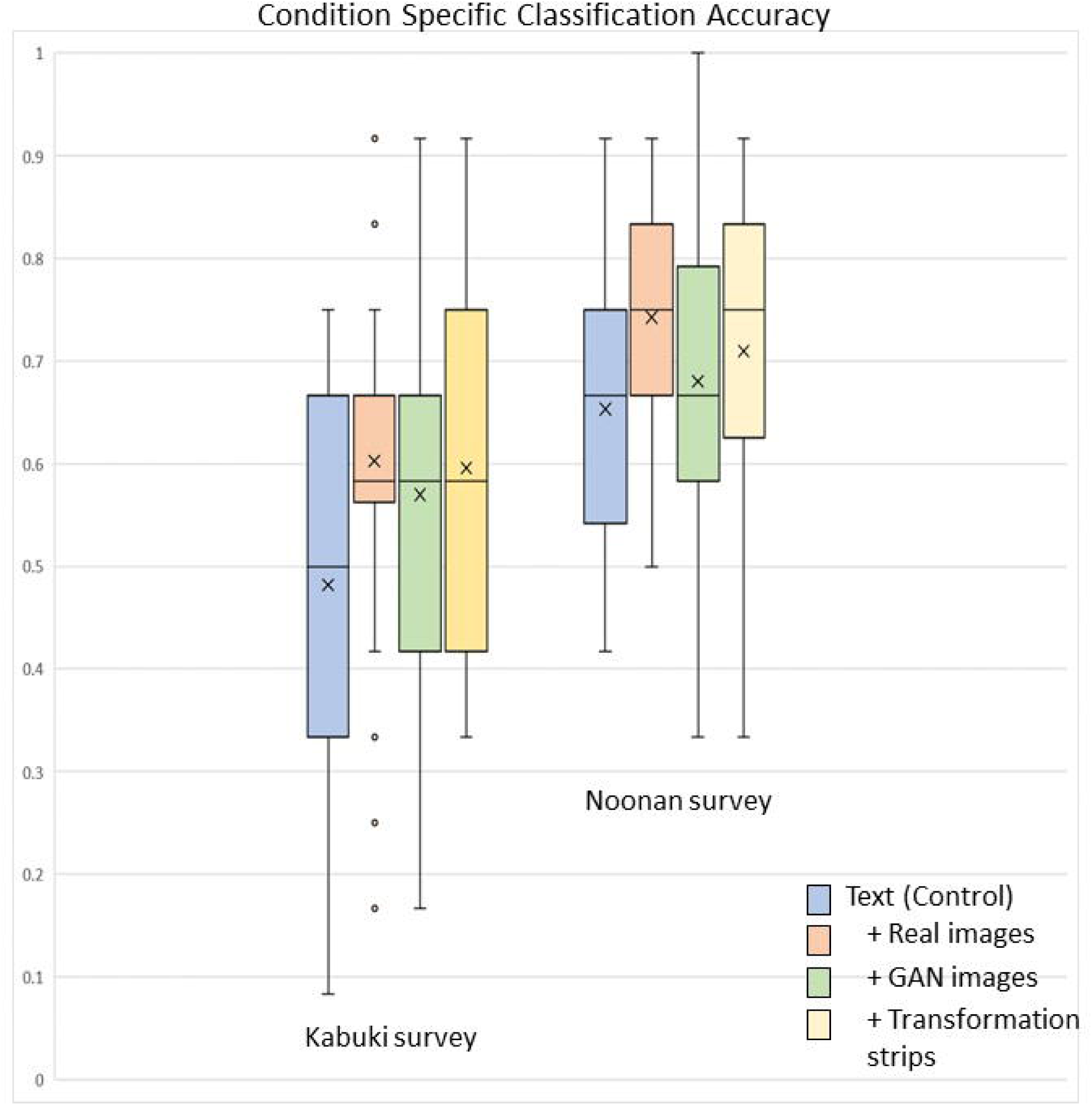
Participant accuracy classifying KS and NS images following educational interventions. Average accuracy (x) increased with all image interventions as compared to text description alone (control). The greatest accuracy increase was observed with KS, where all types of images (real, GAN, and transformation strips) yielded the same median value (line through box plot) and averages nearly 10% higher than text description alone. KS: Kabuki syndrome; NS: Noonan syndrome; GAN: generative adversarial network.

### Attitude and opinion questions

While participants reported a range of confidence levels identifying individuals with the two genetic conditions, participants reported being less confident when assessing KS: 88.7% reported being “not confident” for KS versus NS (51.0%). The proportion of those reporting being confident or somewhat confident when assessing NS was relatively similar from PGY-1 to PGY-3 (range: 44.8-48.3%). All PGY-4 participants in the NS survey (n=4) reported being confident or somewhat confident.

In addition to asking about participants’ overall confidence level in assessing each of the syndromes, we asked participants to rate their confidence level when classifying each individual image. This allowed us to determine whether the educational interventions affected performance as correlated with confidence level (Table 1). When a participant rated their classification answer as highly confident or confident, the average accuracy on these questions was higher than the overall accuracy for the given educational intervention. For example, in the KS survey, the overall accuracy for participants who saw only text descriptions was 48.2%, whereas those classifications that participants rated as being highly confident or confident about had an average accuracy of 62.6%. The average accuracy for classifications that were rated as highly confident or confident increased with any image intervention, ranging from 74% for transformation strips (up from the overall accuracy of 59.6%) to 79.0% for GAN images (up from the overall accuracy of 57.0%) to 81.7% for real images (up from the overall accuracy of 60.3%). Similar trends were observed in the NS survey.

**Table 1.** Impact of self-reported confidence level on accuracy for KS and NS surveys. GAN: generative adversarial network.

|  | Text description<br>only (Control) | Text description<br>+ Real images | Text description<br>+ GAN images | Text description<br>+ Transformation<br>strips |
| --- | --- | --- | --- | --- |
| **Kabuki syndrome images** |  |  |  |  |
| **Overall accuracy** | **48.2%** | **60.3%** | **57.0%** | **59.6%** |
| **Highly confident and confident accuracy** | **62.6%**<br>**(n=46)** | **81.7%**<br>**(n=82)** | **79.0%**<br>**(n=95)** | **74.0%**<br>**(n=79)** |
| **Somewhat confident and not confident accuracy** | **47.2%**<br>**(n=210)** | **52.4%**<br>**(n=229)** | **49.1%**<br>**(n=275)** | **55.7%**<br>**(n=244)** |
| **Noonan syndrome images** |  |  |  |  |
| **Overall accuracy** | **65.3%** | **74.8%** | **68.0%** | **71.0%** |
| **Highly confident and confident accuracy** | **75.9%**<br>**(n=83)** | **84.5%**<br>**(n=116)** | **80.6%**<br>**(n=103)** | **89.4%**<br>**(n=104)** |
| **Somewhat confident and not confident accuracy** | **61.3%**<br>**(n=217)** | **67.72%**<br>**(n=158)** | **61.4%**<br>**(n=197)** | **63.1%**<br>**(n=244)** |

At the end of the survey, participants were asked to rate the usefulness of the educational intervention. Approximately, 60% of participants who received text only description found the text description to be useful for both KS and NS. Interestingly, the reported usefulness of the text increased when coupled with any image type (Table 2). For example, the text descriptions were considered useful by more participants when coupled with real KS (76.9%) and NS (78.3%) images. While over 90% of participants rated both KS and NS images as useful, they rated GAN images and transformation images as less useful. For example, 96.2% of participants seeing real KS images rated them useful, whereas, only 64.5% (GAN) and 74.1% (transformation) images were rated as useful. We note that accuracy performance for KS surveys was 57.0-60.3% for any image intervention coupled with text, as compared to 48.2% for text only description (Figure 1). Additionally, participants were aware of whether images were real or AI-generated, which have may affected how they rated perceived usefulness.

**Table 2.** Usefulness of text descriptions and image intervention in perceived KS and NS survey performance. GAN: generative adversarial network.

|  | Kabuki syndrome |  |  |  | Noonan syndrome |  |  |  |
| --- | --- | --- | --- | --- | --- | --- | --- | --- |
|  | Text description only (Control) | Text description + Real images | Text description + GAN images | Text description + Transformation image strips | Text description only (Control) | Text description + Real images | Text description + GAN images | Text description + Transformation image strips |
| Participants that found text description useful | 59.1% | 76.9% | 58.1% | 66.7% | 60% | 78.3% | 62.5% | 58.6% |
| Participants that found image intervention useful | N/A | 96.2% | 64.5% | 74.1% | N/A | 91.3% | 79.2% | 75.9% |
| Text only intervention participants that would have found images to be useful | 81.8% | N/A | N/A | N/A | 100% | N/A | N/A | N/A |

Participants were asked both prior to the educational intervention and at the end of the survey about their opinion as to which facial features were important (Figure 2). Over half of participants in the KS survey reported they were unsure about important diagnostic facial features for that syndrome, and 24.0% of NS survey participants were unsure. At the conclusion of the survey, the number of participants who reported being unsure dropped to less than 10% for both conditions. Regardless of educational intervention, the number who selected the appropriate (based on OMIM annotations) specific facial features increased. For KS, the greatest changes were observed in the correct selection of eyes, ears, and nose, which was overall greater in participants that received image interventions (File 7). The greatest changes between pre– and post-survey for NS was an increase in the number of participants correctly reporting eyes and ears as important features, and a decrease in the number of participants reporting mouth as an important feature (File 8).

**Figure 2.**
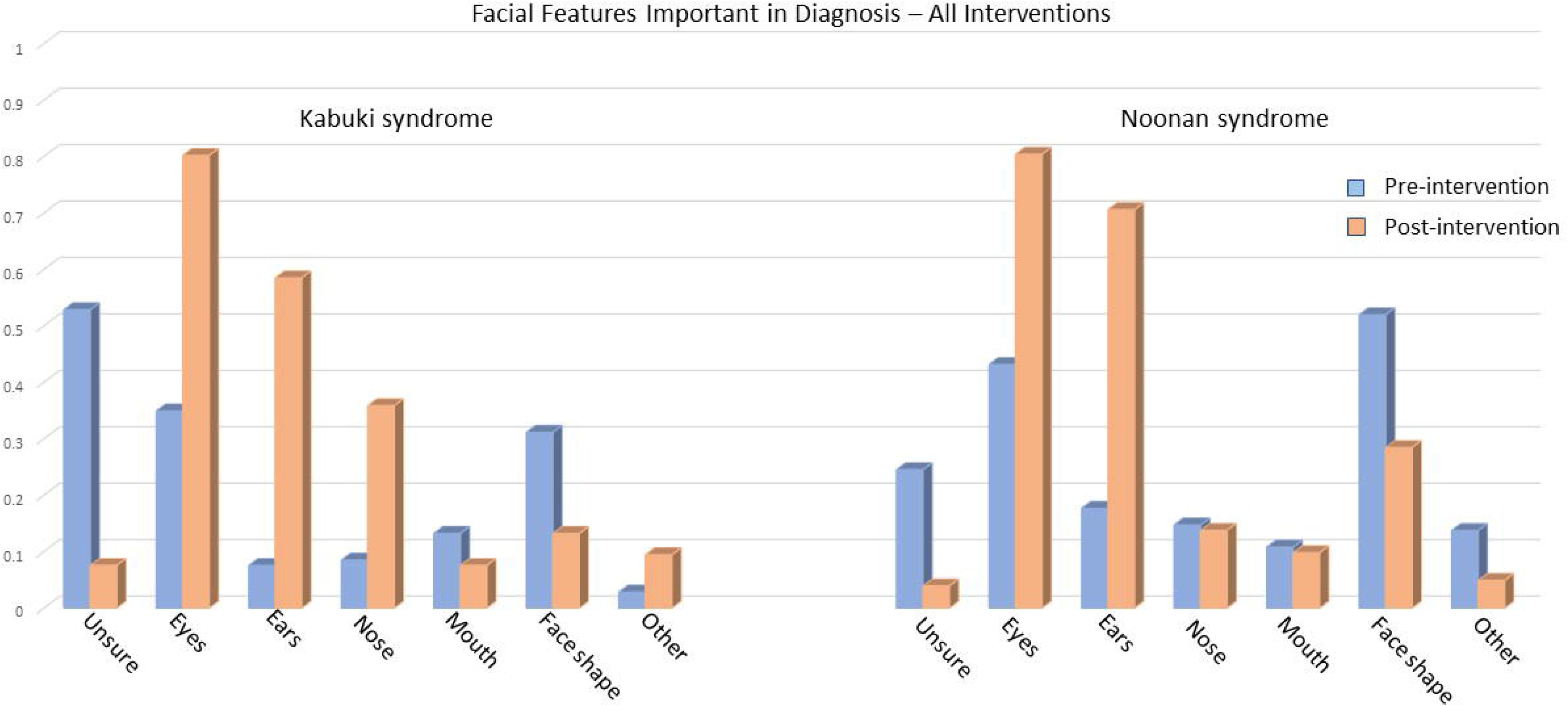
Self-reported facial features important in diagnosis, pre– and post-intervention. All interventions increased awareness of important facial features in both KS and NS, a decrease in unsure response and an increase in typical dysmorphic features (KS: eyes, ears, nose and NS: eyes and ears). KS: Kabuki syndrome; NS: Noonan syndrome

To better understand the perceived impact of age, ancestry, and gender on syndromic facial features, we asked participants to report how much each factor influences the facial features of each syndrome. Prior to providing the education interventions, 73.1-78.3% of KS and 33.3-44.1% of NS survey respondents (depending on the survey) reported being unsure of the influence of these factors on syndromic facial features (File 9). Nearly 50% of NS survey participants reported prior to the educational intervention that age has some or great influence on facial features. Post survey, 100% of participants reported an opinion about the influence of these 3 factors on facial features (Files 10 and 11). Interestingly, for both the KS and NS surveys, those receiving only the text intervention, reported gender as having more influence on facial features, 20.6% and 23.1%, respectively than the average of those that received text and any image type. While other difference may exist with gender and ancestry based on educational intervention, it would require a greater sample size to confirm.

## Discussion

There is understandable excitement around recent developments in AI. This is tempered by many concerns, including that careful testing needs to be done to ensure that the benefits of AI outweigh risks.

In our study, we tested two key measures: participants’ opinions and self-reported confidence as well as actual improvement in participants’ knowledge (accuracy). Prior to the educational intervention, 53% and 24% of the KS and NS survey participants respectively reported that they were unsure about the important diagnostic facial features for that syndrome. At the end of the survey, the percentage of participant who were unsure dropped to 8% and 4% for KS and NS respectively (Figure 2). For KS, a syndrome less familiar to the pediatric residents, accuracy improved by 12% when real images were added to the control text description intervention. An improvement in accuracy was also observed (of 9% and 11%, respectively) when GAN images and GAN transformation strips were added (Figure 1).

Although generative AI did not outperform real images as an educational tool, our results imply that generative AI may be a helpful adjunct in medical genetics education to expose trainees to more images. Advantages of using generative AI includes that, after initial training, many images can be made quickly, and images can be selected to ensure representations of diverse individuals. Related to this, it is critical to emphasize that further study is needed to ensure that AI tools work equitably across different populations.^25, 26^ Generative AI can also help address issues of privacy and data sharing by avoiding the use of actual patient images. Finally, our (and other) results also show that realistic images can be generated with relatively small datasets such as may be available in the context of rare diseases.^8, 21, 22^

Our study has limitations. We only examined two different conditions, which were selected as representing genetic disorders with which pediatric residents would likely have different familiarity levels. The study also had a small number of participants, and we tested our methods through an online survey, versus trying other teaching techniques. Further testing to include more conditions, more participants and learning modalities, and different types of data, could help provide a broader sense of the usefulness (or not) of these approaches. Given our promising results with the current study, another possible approach is a user-interactive platform, where participants could generate far more than 5 images, as well as collect user feedback.

## Conclusion

In conclusion, we feel that our methods do not imply that generative AI could somehow replace traditional teaching methods but could be an opportunity for AI–human collaboration to enhance genetics education, especially as relates to rare syndromes. Though currently the topic of a great deal of conversation, AI might best be viewed as another means to reach different styles of learners and provide genetics content in new ways. These approaches can be particularly useful in programs with little contact with medical geneticists, or where a dedicated genetics elective may not be available.

## Data Availability

All data produced in the present study are available in this paper, on our GitHub site, or upon reasonable request to the authors.

https://github.com/datduong/stylegan3-syndromic-faces

## Acknowledgements

This research was supported in part by the Intramural Research Program of the National Human Genome Research Institute, National Institutes of Health. This work utilized the computational resources of the NIH HPC Biowulf cluster.

## FIG

## References

1 Ledgister Hanchard, S. E., et al. Scoping review and classification of deep learning in medical genetics. Genet Med 24, 1593–1603, doi:10.1016/j.gim.2022.04.025 (2022).

2 Porras, A. R., Rosenbaum, K., Tor-Diez, C., Summar, M. & Linguraru, M. G. Development and evaluation of a machine learning-based point-of-care screening tool for genetic syndromes in children: a multinational retrospective study. Lancet Digit Health 3, e635–e643, doi:10.1016/S2589-7500(21)00137-0 (2021).

3 Luo, R., Sedlazeck, F. J., Lam, T. W. & Schatz, M. C. A multi-task convolutional deep neural network for variant calling in single molecule sequencing. Nat Commun 10, 998, doi:10.1038/s41467-019-09025-z (2019).

4 Ledgister Hanchard, S. E., Dwyer, M.C., Liu, S., Hu, P., Tekendo-Ngongang, C., Waikel, R.L., Duong D. Solomon, B.D. Scoping review and classification of deep learning in medical genetics. Genet Med (2022).

5 Hsieh, T. C. et al. GestaltMatcher facilitates rare disease matching using facial phenotype descriptors. Nat Genet 54, 349–357, doi:10.1038/s41588-021-01010-x (2022).

6 Karras, T. et al. Training generative adversarial networks with limited data. arXiv preprint arXiv:2006.06676 (2020).

7 Jeong, J. J. et al. Systematic Review of Generative Adversarial Networks (GANs) for Medical Image Classification and Segmentation. J Digit Imaging 35, 137–152, doi:10.1007/s10278-021-00556-w (2022).

8 Malechka, V. V. et al. Investigating Determinants and Evaluating Deep Learning Training Approaches for Visual Acuity in Foveal Hypoplasia. Ophthalmol Sci 3, 100225, doi:10.1016/j.xops.2022.100225 (2023).

9 Chan, K. S. & Zary, N. Applications and Challenges of Implementing Artificial Intelligence in Medical Education: Integrative Review. JMIR Med Educ 5, e13930, doi:10.2196/13930 (2019).

10 Arora, A. Disrupting clinical education: Using artificial intelligence to create training material. Clin Teach 17, 357–359, doi:10.1111/tct.13177 (2020).

11 Chen, J. S. et al. Deepfakes in Ophthalmology: Applications and Realism of Synthetic Retinal Images from Generative Adversarial Networks. Ophthalmol Sci 1, 100079, doi:10.1016/j.xops.2021.100079 (2021).

12 Solomon, B. D. Can artificial intelligence save medical genetics? Am J Med Genet A, doi:10.1002/ajmg.a.62538 (2021).

13 Gonzaludo, N., Belmont, J. W., Gainullin, V. G. & Taft, R. J. Estimating the burden and economic impact of pediatric genetic disease. Genet Med 21, 1781–1789, doi:10.1038/s41436-018-0398-5 (2019).

14 Ferreira, C. R. The burden of rare diseases. Am J Med Genet A 179, 885–892, doi:10.1002/ajmg.a.61124 (2019).

15 Jenkins, B. D., et al. The 2019 US medical genetics workforce: a focus on clinical genetics. Genet Med, doi:10.1038/s41436-021-01162-5 (2021).

16 Rubanovich, C. K., Cheung, C., Mandel, J. & Bloss, C. S. Physician preparedness for big genomic data: a review of genomic medicine education initiatives in the United States. Hum Mol Genet 27, R250–r258, doi:10.1093/hmg/ddy170 (2018).

17 Forsyth, R. et al. A structured genetics rotation for pediatric residents: an important educational opportunity. Genet Med 22, 793–796, doi:10.1038/s41436-019-0723-7 (2020).

18 Gates, R. W., Hudgins, L. & Huffman, L. C. Medical genetics education for pediatrics residents: A brief report. Genet Med 24, 2408–2412, doi:10.1016/j.gim.2022.08.003 (2022).

19 Carroll, J. C. et al. The Gene Messenger Impact Project: An Innovative Genetics Continuing Education Strategy for Primary Care Providers. J Contin Educ Health Prof 36, 178–185, doi:10.1097/CEH.0000000000000079 (2016).

20 Kimmerle, J., Timm, J., Festl-Wietek, T., Cress, U. & Herrmann-Werner, A. Medical Students’ Attitudes toward AI in Medicine and their Expectations for Medical Education. medRxiv, 2023.2007.2019.23292877, doi:10.1101/2023.07.19.23292877 (2023).

21 Duong, D., Waikel, R. L., Hu, P., Tekendo-Ngongang, C. & Solomon, B. D. Neural network classifiers for images of genetic conditions with cutaneous manifestations. HGG Adv 3, 100053, doi:10.1016/j.xhgg.2021.100053 (2022).

22 Duong, D. et al. Neural Networks for Classification and Image Generation of Aging in Genetic Syndromes. Front Genet 13, 864092, doi:10.3389/fgene.2022.864092 (2022).

23 Niikawa, N. et al. Kabuki make-up (Niikawa-Kuroki) syndrome: a study of 62 patients. Am J Med Genet 31, 565–589, doi:10.1002/ajmg.1320310312 (1988).

24 Romano, A. A. et al. Noonan syndrome: clinical features, diagnosis, and management guidelines. Pediatrics 126, 746–759, doi:10.1542/peds.2009-3207 (2010).

25 Tekendo-Ngongang, C. et al. Rubinstein-Taybi syndrome in diverse populations. Am J Med Genet A 182, 2939–2950, doi:10.1002/ajmg.a.61888 (2020).

26 Solomon, B. D. et al. Perspectives on the future of dysmorphology. Am J Med Genet A, doi:10.1002/ajmg.a.63060 (2022).

