## Supplementary figures and images for "Generative Methods for Pediatric Genetics Education"

### Supplemental File 2

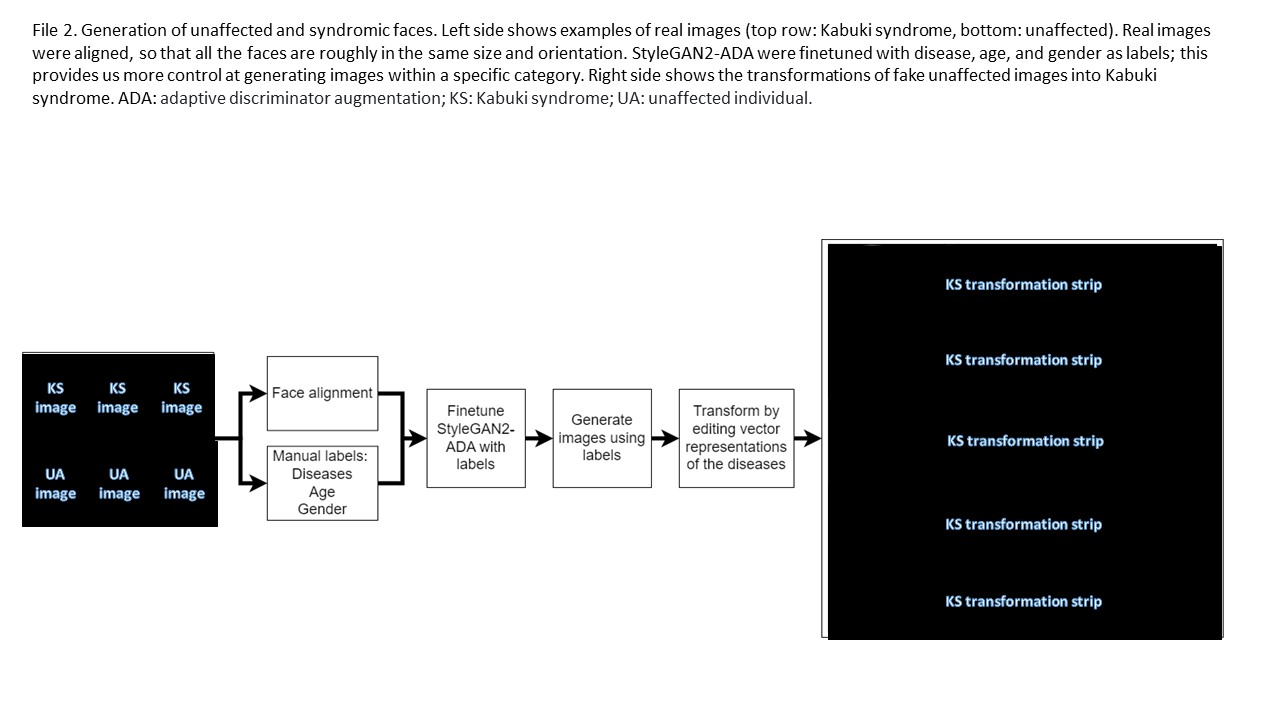

### Supplemental File 3

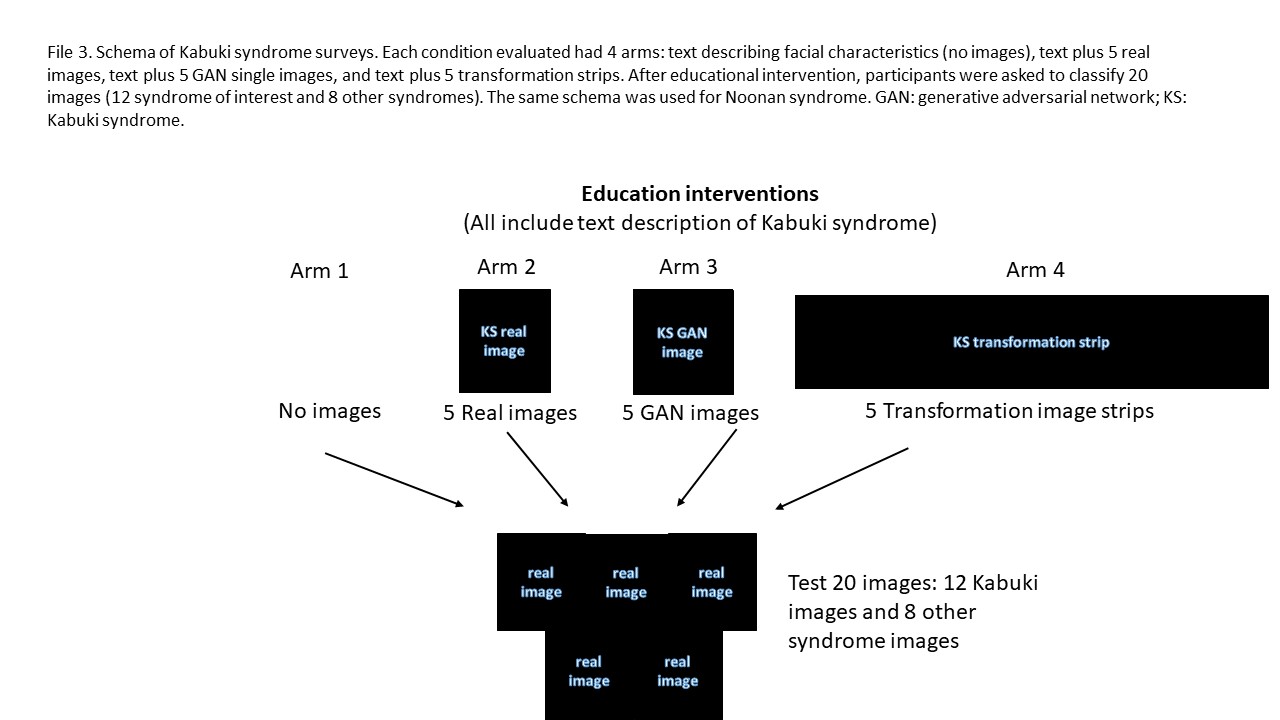

### Supplemental File 4

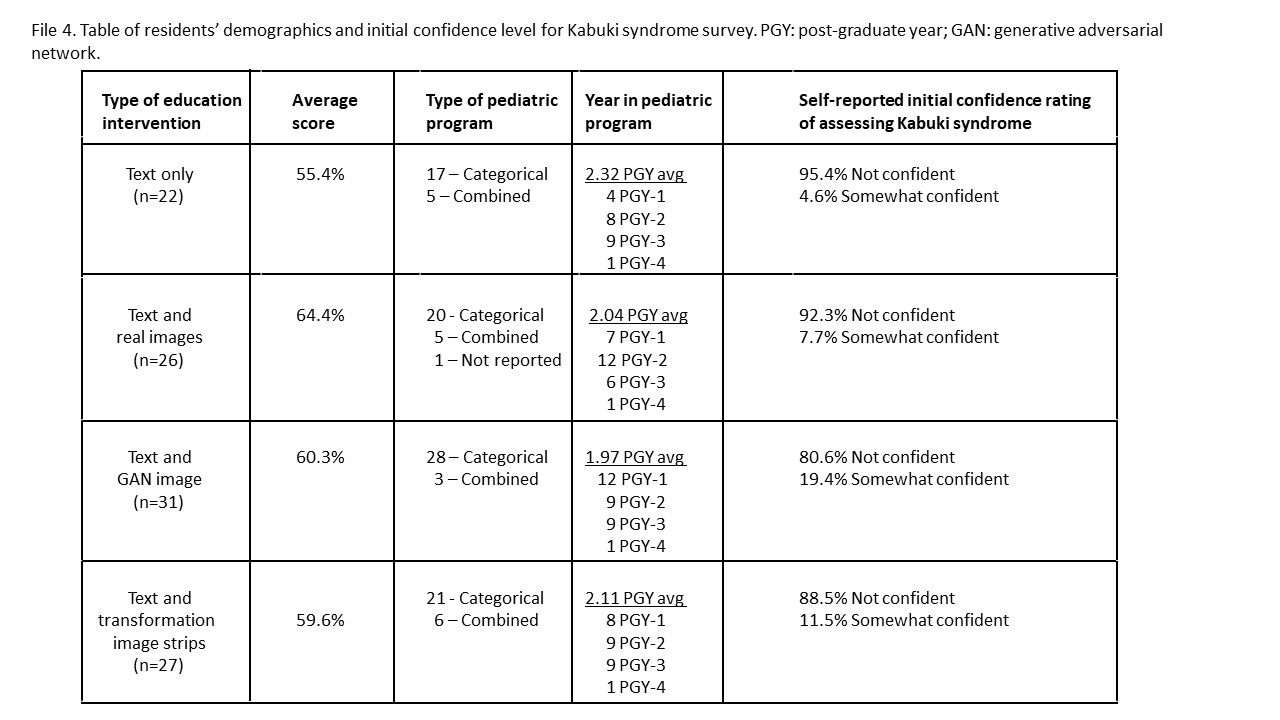

### Supplemental File 5

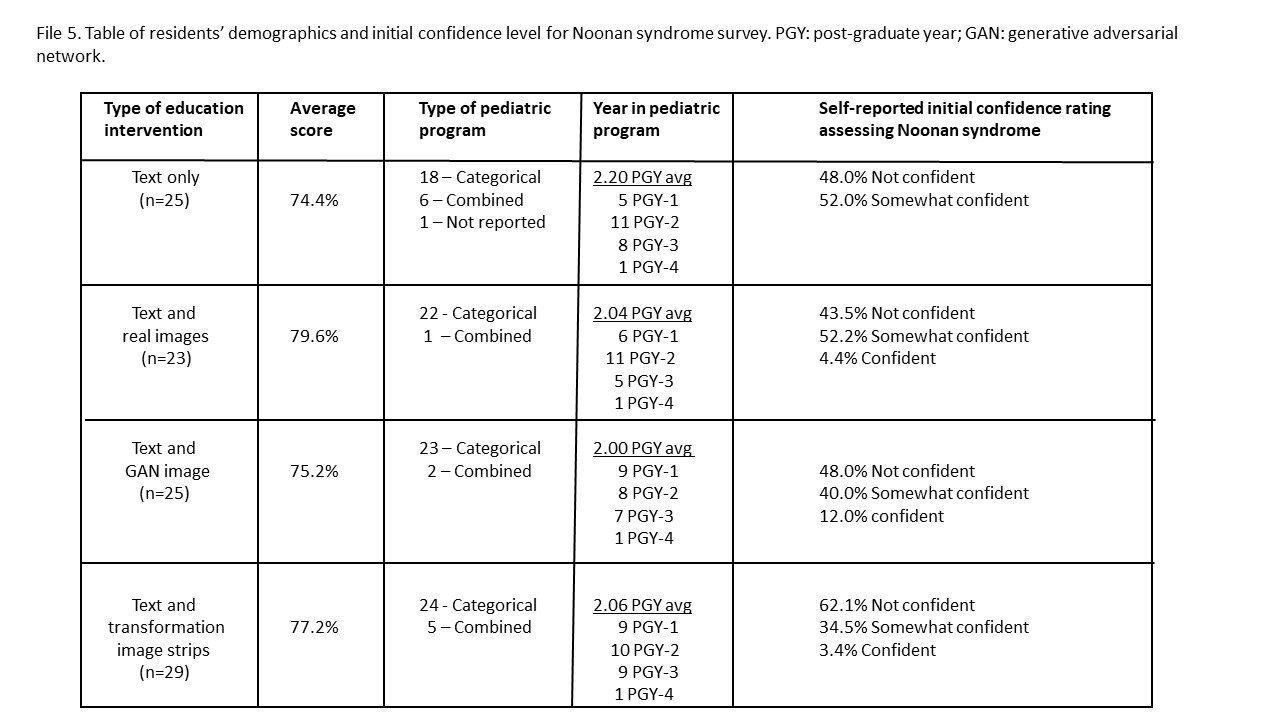

### Supplemental File 6

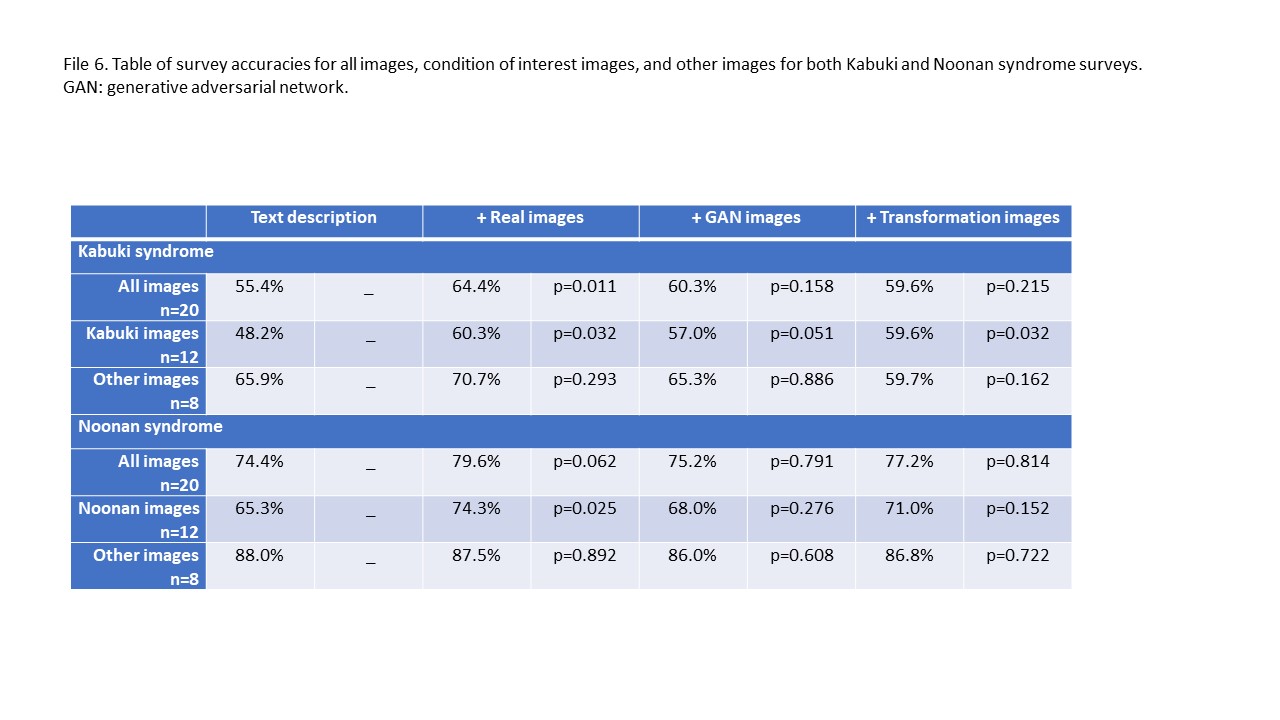

### Supplemental File 7

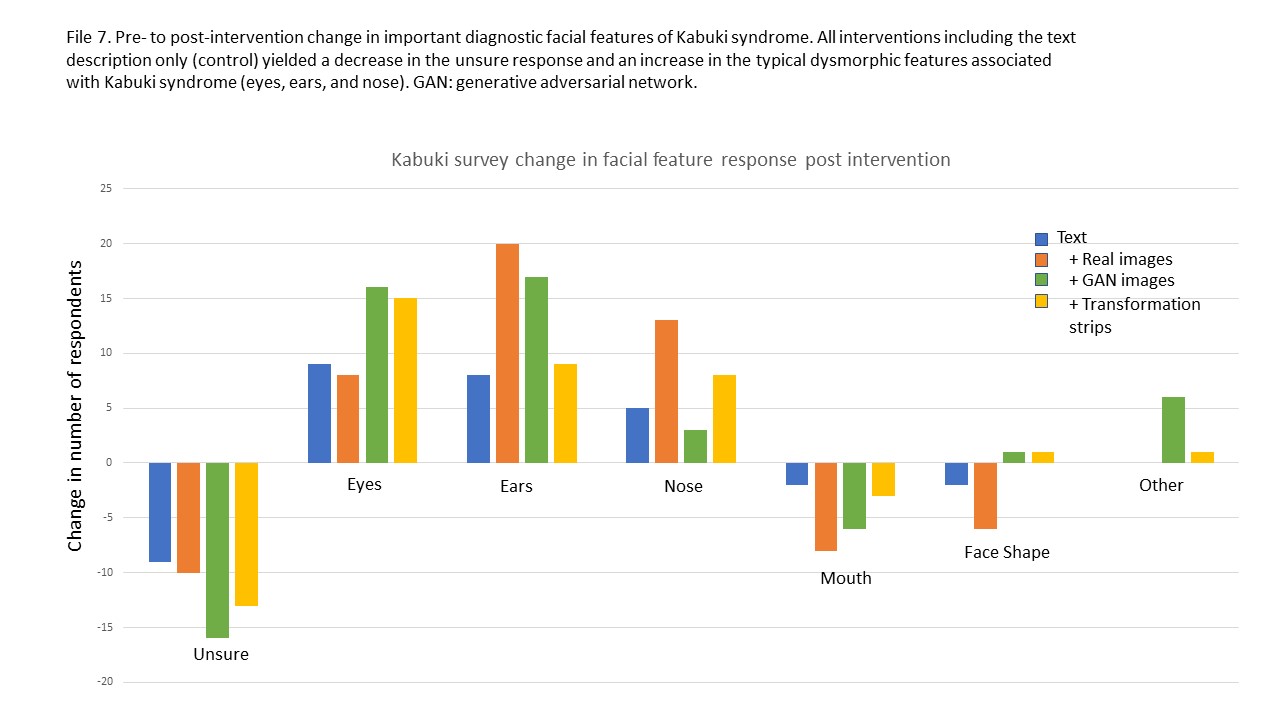

### Supplemental File 8

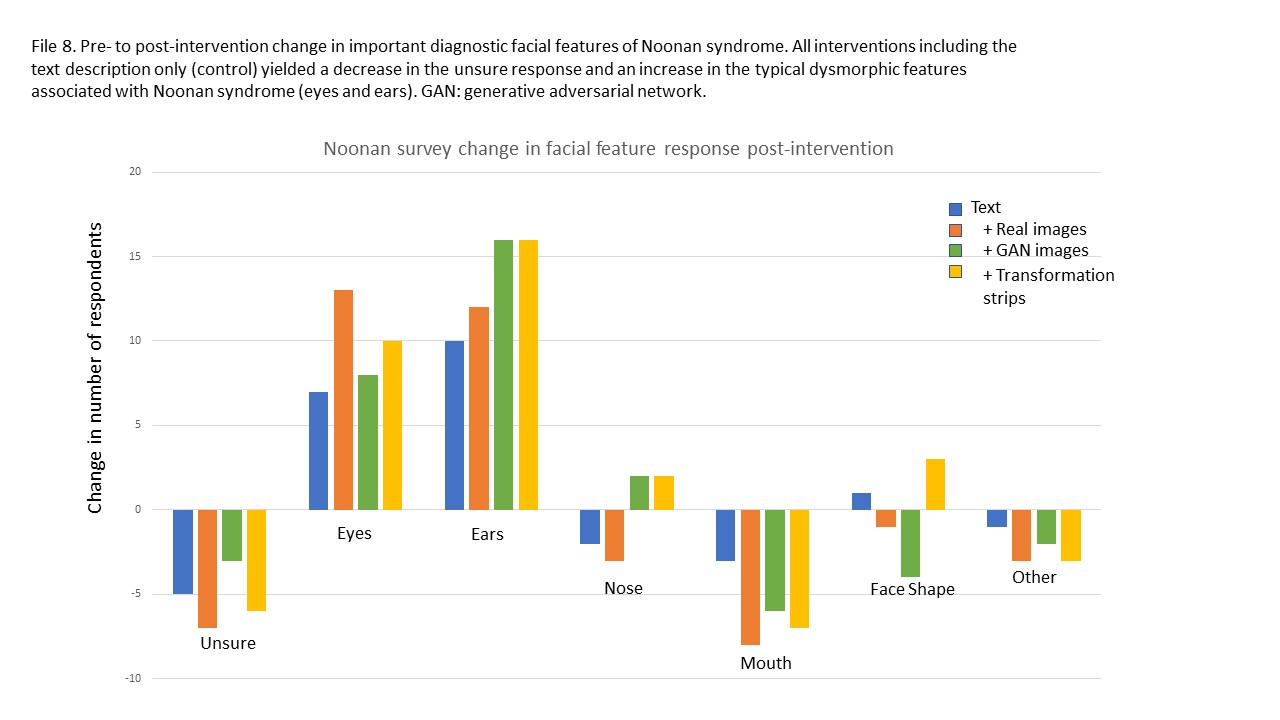

### Supplemental File 9

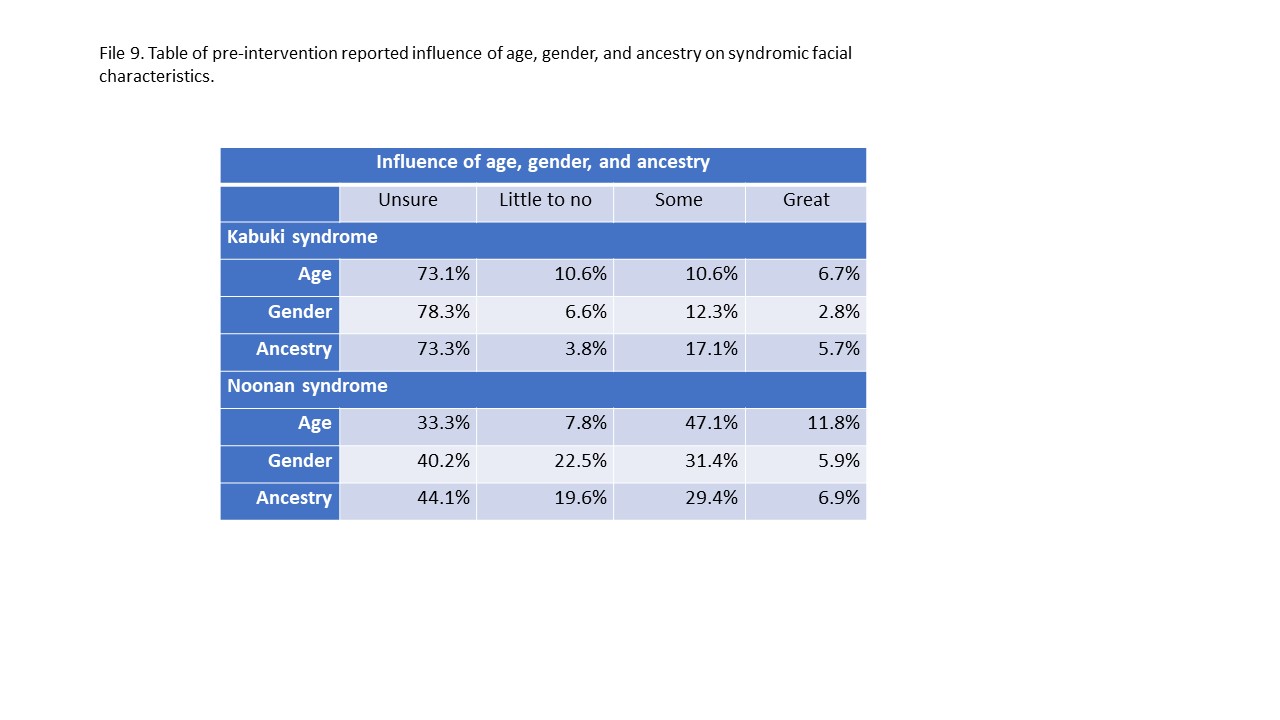

### Supplemental File 10

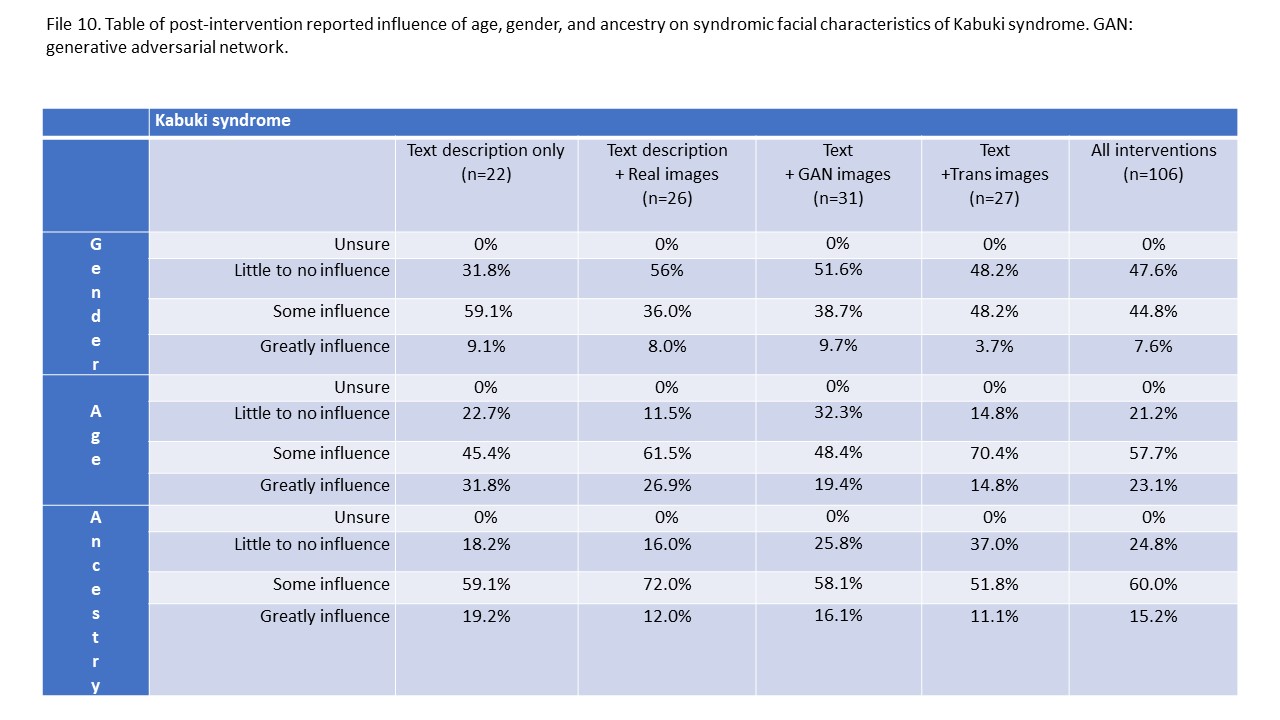

### Supplemental File 11

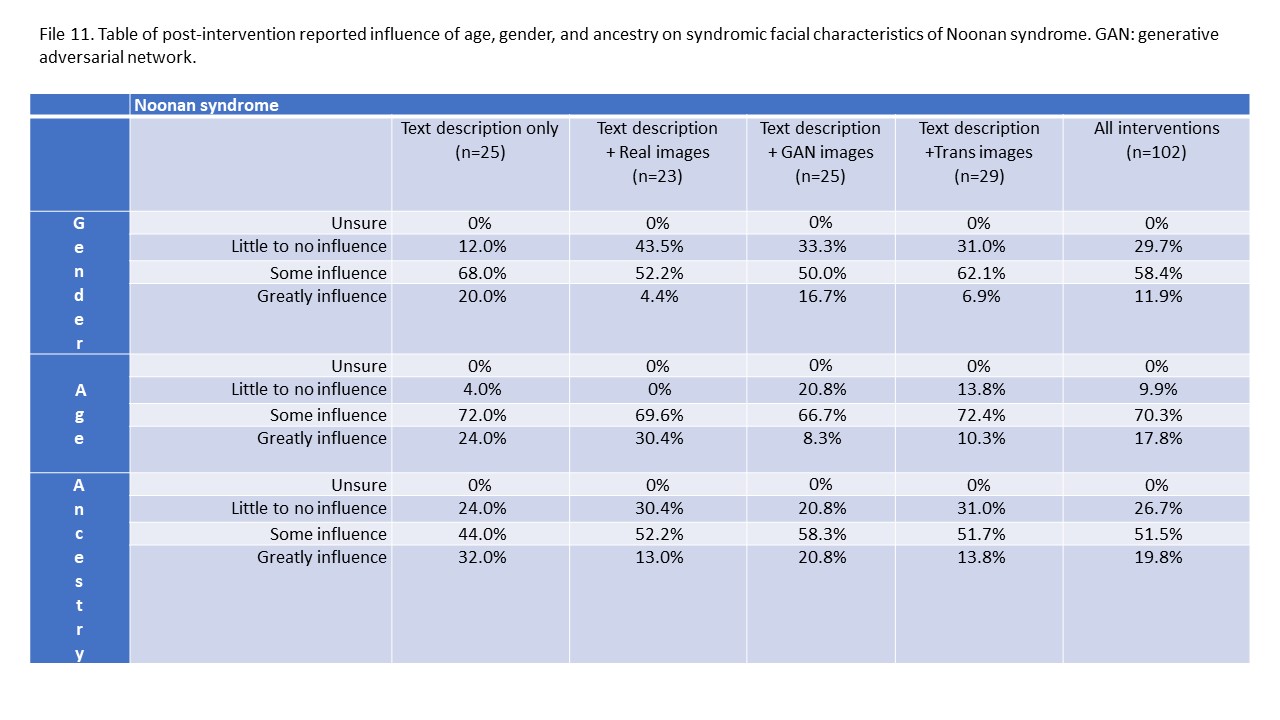
